# Digital healthcare reform and reduction of duplicate examinations and tests: a study of physicians’ behavior on mutual recognition in response to clinical specialized demands and information intervention form a national pilot province in China

**DOI:** 10.1101/2025.01.26.25321151

**Authors:** Chao Song, Xinmian Huang, Shasha Qian, Chaoyun Yuan, Shuning Liu, Jun Zhou

**Affiliations:** Department of Medical Affairs and Zhejiang Center of Clinical Laboratories, Zhejiang Provincial People’s Hospital, People’s Hospital of Hangzhou Medical College, Hangzhou, China.; Department of Urology and Medical Affairs, Zhejiang Provincial People’s Hospital, People’s Hospital of Hangzhou Medical College, Hangzhou, China.; Department of Medical Affairs, Zhejiang Provincial People’s Hospital, People’s Hospital of Hangzhou Medical College, Hangzhou, China.; Department of Information Center, Zhejiang Provincial People’s Hospital, People’s Hospital of Hangzhou Medical College, Hangzhou, China.; Department of International and Domestic Cooperation, Zhejiang Provincial People’s Hospital, People’s Hospital of Hangzhou Medical College, Hangzhou, China.; Department of Laboratory Medicine, Hangzhou Children’s Hospital, Hangzhou, China.

**Keywords:** Mutual Recognition, Hospital Management, Clinical Specialties, Indicator Management Method, Information Intervention, Medical Insurance

## Abstract

**Background:** To reduce duplicate medical exams and tests for patients across different hospitals and alleviate their financial burden, Zhejiang, with the capital Hangzhou, has launched a digital healthcare reform named “Zhejiang Medical Mutual Recognition” from 2021 and become a pilot province in China. This digital healthcare reform policy has begun to be gradually implemented nationwide. This study aims to evaluate and analyze the differences in physicians’ behavior during the mutual recognition process across different types of hospitals, clinical specialties, and information intervention strategies, in order to provide suggestions and assistance for the optimization and improvement of nationwide mutual recognition policies.

**Methods:** This is a one-year multicenter study involving eight top-tier hospitals in Hangzhou, China, covering various types of hospitals, including general hospitals, traditional chinese medicine hospitals, integrated chinese and western medicine hospitals, and specialized hospitals. A set of recognition indicators, such as recognition proportion, cross-hospital recognition rate, were designed to evaluate physicians’ behaviors, providing a multi-dimensional perspective. Hospitals were grouped and compared based on their characteristics of their clinical specialties. The key recognition indicators among different hospitals, different specialties, and the same specialties in different hospitals were compared. The information intervention strategies were implemented in 3 hospitals to reduce the overlooked access rate and improve recognition rates through the method of information system restrictions. The remaining five hospitals, which did not implement these specific information interventions, served as the control group.

**Results:** The traditional chinese medicine demonstrated a low cross-hospital precision delivery rate but a high rate of recognizing reports from other hospitals, contrasting with pediatrics. There were significant differences in recognition indicators among different clinical specialties. The total recognition proportion for the traditional chinese medicine group and the pediatrics group were 51.18% and 9.95%, respectively. The differences were not significant among the same clinical specialties in different hospitals, however, some recognition indicators were also noticeably affected by the mutual recognition management strategies of the affiliated hospitals. Information intervention in certain hospitals significantly reduced the overlooked access rate; information intervention had increased the workload for physicians in accessing duplicate reports, mainly stemming from the repetitive process of accessing reports from their own hospital both locally and on the platform, without significant affect on the main recognition indicators across hospitals.

**Conclusion:** The recognition indicators designed in this study can effectively assess and provide decision support for mutual recognitio management. Although the treatment characteristics of clinical specialties are factors affecting mutual recognition, the management strategies implemented by hospitals can also significantly change mutual recognition. Hospital management strategies, such as information interventions, need to strike a balance between mutual recognition work and the workload of physicians accessing duplicate reports.

## Introduction

In the context of healthcare reform in China,[1] the mutual recognition of medical examination and test reports (MR) has emerged as a pivotal strategy to enhance the efficiency of the healthcare system and reduce the burden on patients.[2,3] The Diagnosis-Related Groups (DRG) payment method controls medical expenses by setting predetermined cost standards, encouraging hospitals to use resources efficiently.[4–6] The MR reduces unnecessary duplicate examinations and tests, thereby lowering costs. When integrated with the DRG payment method, it aids in managing medical insurance expenditures and enhancing the efficiency of medical services.[7] The MR had significantly reduced the unnecessary repetitive examinations and tests when patients seek treatment across different hospitals in the past three years, enhanced the efficiency of tiered diagnosis and treatment, and also markedly saved on the expenditure of medical insurance funds.

Zhejiang Province, with its provincial capital Hangzhou, is situated in the eastern coastal region of China within the Yangtze River Economic Belt and is acknowledged as a leading province in MR, serving as a model for the advancement of Shanghai, Jiangsu Province, and Fujian Province.[8] Zhejiang Province has taken the lead in establishing the “Zhejiang Medical Mutual Recognition” platform (MR platform) in 2021[9], which is dedicated to breaking down barriers in medical data and achieving seamless MR and efficient sharing of examination and test results across diverse medical institutions. The MR platform has become a key focus for enhancing medical efficiency, optimizing resource allocation, and alleviating the economic burden on patients.[10] On the government website of Zhejiang Province in China, as of 2022, the MR platform had cumulatively recognized various items 16.81 million, directly saving medical expenses of 712 million RMB, with residents making over 70,000 self-service inquiries daily.[11]

The MR platform is built upon a province-wide integrated intelligent public data architecture, deeply integrating medical big data resources and seamlessly connecting with various departmental medical information systems. It activates the potential of medical data elements and reshapes the value ecosystem of medical and health services. Through digital empowerment, the platform standardizes diagnostic and treatment processes in all aspects, strengthens mechanisms for rational examinations, enhances supervision and management efficiency, and significantly improves the patient’s medical experience. Since the MR platform’s launch three years ago, by the end of 2024, it has been deeply integrated into clinical practice. Physicians have become proficient in using the platform to access patients’ previous examination and test reports, which has significantly reduced diagnostic cycles, markedly improving the efficiency and quality of medical services, and effectively reducing the burden on patients. It has also accumulated experience in MR management for medical institutions and prepared for the reform of medical insurance fund payments.

However, it cannot be ignored that the diversity of hospital management and the complexity of clinical specialty characteristics are intertwined, forming a multifaceted network of factors affecting the effectiveness of MR. The underlying mechanisms of this network remain to be deeply explored. This gap hampers the precise optimization and widespread promotion of MR policies, necessitating systematic research to address it. In light of this, our study is based on the MR platform, deeply analyzing the interactive logic of hospital management and clinical specialty characteristics. We aim to customize strategies for the refined hospital management of recognition and provide a basis for the construction of a national MR network and the assessment of physicians’ behaviors.

## Methods

### Participants and data collection

This study was a multicenter analysis, including various types of hospitals across China, all of which were large tertiary hospitals (grade A hospitals), and were concentrated in the provincial capital city of Hangzhou. A total of eight hospitals were included in the focal study, representing a diverse array of medical institutions. These hospitals were selected based on the scale of their operations, with each exceeding an annual medical revenue of 1.6 billion RMB (approximately 225 million USD) in 2023.[12] The participants were composed of four general hospitals (designated as hospital A, hospital B, hospital C, and hospital D), a hospital specializing in traditional chinese medicine(TCM) (designated as hospital E), an integrated hospital combining chinese and western medical practices (designated as hospital F), and two specialized hospitals (designated as hospital G and hospital H). The selection criteria ensured a comprehensive evaluation across various healthcare delivery models.

The physician behaviors of eight hospital were recorded by the MR platform, which was accessible to hospitals. A complete statistical period from October 1, 2023, to September 30, 2024, was selected to ensure a comprehensive reflection of physician behaviors, mitigating the impact of seasonal or short-term fluctuations and enhancing the representativeness and stability.

All the data were subjected to anonymization. This process involved the careful exclusion of any sensitive information that could compromise individual privacy or reveal clinical symptoms, thus aligning with regulatory compliance and prioritizing patient confidentiality. All the data did not contain information that could identify individual participants during or after data collection.

### Mutual recognition workflow

The recognition and the specific actions taken by physicians at each step are shown in the workflow (Fig 1). The workflow begins with a physician’s intention to issue exam or test items order. The MR platform is then queried to search for the existence of recently completed reports for the same items in the patient’s medical records from other hospitals. If no report is found, the item order can be directly sent and executed by the local hospital. However, if there is a recently conducted report of the same item exists, the platform will deliver the reports to the local medical system and remind the physician to access the delivered reports. Upon accessing the reports, the physician should make a decision on recognition. If the decision is non-recognition, the item order issued by physician will be executed by the local hospital. Conversely, a recognition decision will lead to the cancellation of the issued item order, and the recognized reports will be downloaded to the local medical system. If the physician consistently overlooks the delivered reports, the item order can also be executed by the local hospital.

**Fig 1.**
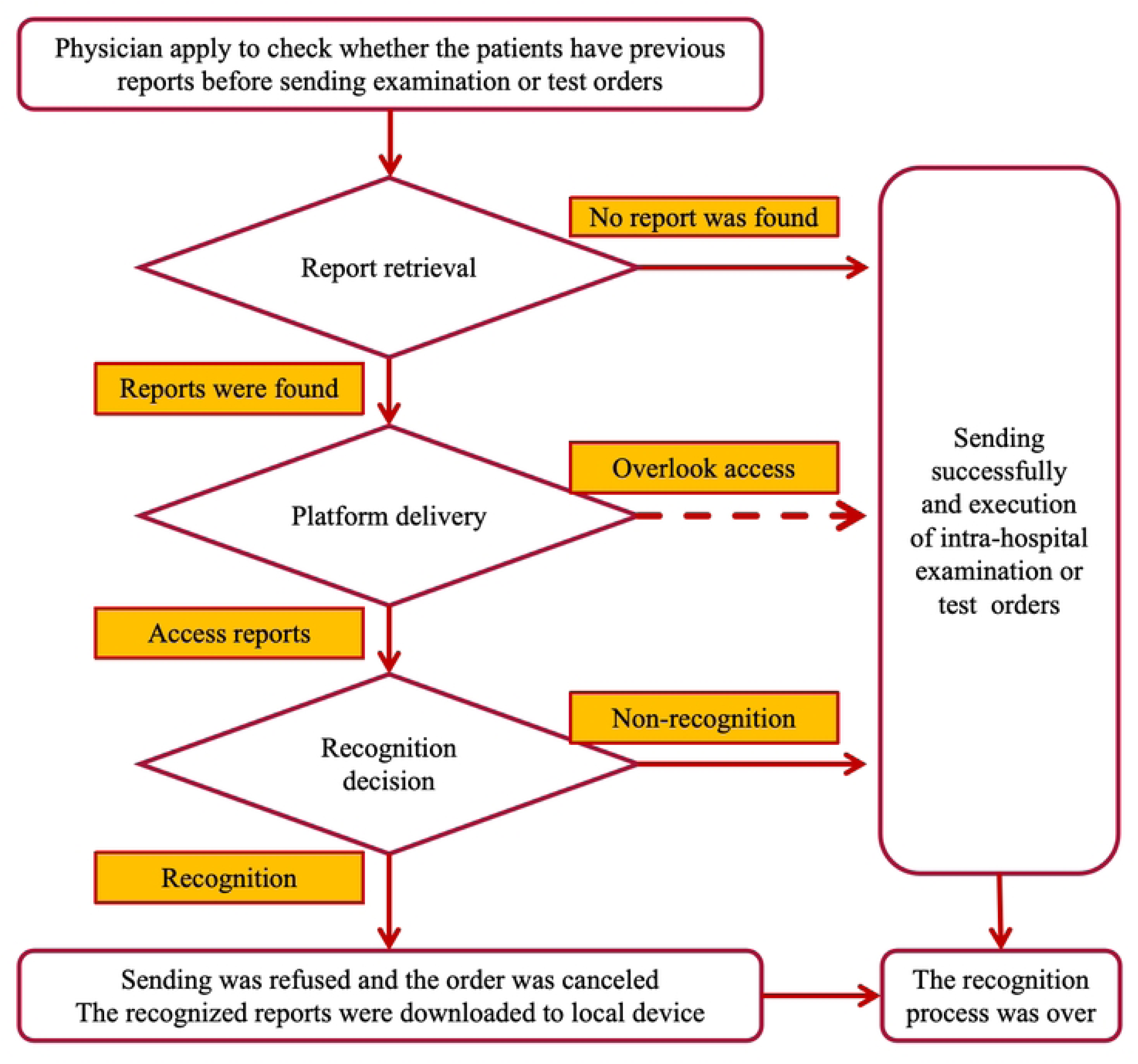
The workflow of the recognition process in the MR platforr

### Recognition indicators

A set of recognition indicators were designed to analyze the activities of hospitals, clinical specialties, and physicians on the MR platform. The main recognition indicators were designed based on the frequency of platform delivery, access reports, recognition decision, providing a quantitative assessment of recognition. The definitions and connotations of each recognition indicator were detailed as flowers.

**Total Recognition Proportion(TRP):** This indicator showed the percentage of recognized items to the overall accessed items.

**Total Recognition Rate(TRR)**:This indicator showed the percentage of recognized items to the delivered items.

**Precision Delivery Rate(PDR):** This indicator showed the percentage of the precision delivered reports to the total number of delivered reports within a standard period, reflecting how well a patient’s previous medical reports align with their current medical needs.

**Access Rate(AR):** This indicator showed the percentage of the accessed items to the total number of precision delivered items, indicating the effectiveness of physicians’ engagement with the delivered items.

**Overlooked Access Rate(OAR)**: This indicator showed the percentage of the overlooked items to the delivered items, highlighting instanced where physicians might overlook or choose not to engage with exams and tests.The Access Rate and the Overlooked Access Rate sum up to 100%.

**Cross-Hospital Access Rate(CHAR):** This indicator showed the percentage of the accessed items to the precision delivered items from other hospital, indicating the effectiveness of physicians’ engagement with the other hospitals’ delivered items.

**Cross-Hospital Recognition Proportion(CHRP):**This indicator showed the percentage of recognized items to the accessed items from other hospitals.

**Cross-Hospital Recognition Rate(CHRR)**: This indicator showed the percentage of the recognized items in total delivered items from other hospitals, measuring the effectiveness of cross-hospital recognition.

**Intra-Hospital Recognition Proportion(IHRP):**This indicator showed the percentage of recognized items to the accessed items from the hospital where the physician was based.

**Radiological Exam Fees Proportion(REFP):** This represented the share of cost savings that came from recognizing radiological exam items in the total cost of all recognized items, revealing the efficiency of radiological exam resources used in the recognition.

### Groups and Comparisons

This analysis involved a comparison of various recognition indicators across 8 hospitals. The comparisons at the clinical specialty level began by categorizing data into four major groups: internal medicine, surgery, TCM, and pediatrics. Data from outpatient departments or units named under internal medicine were allocated to the internal medicine group, and similarly, data named under surgery were allocated to the surgery group, traditional chinese medicine to the traditional chinese medicine group, and pediatrics to the pediatrics group. To further delineated the behavioral differences among clinical specialties, the surgery and internal medicine groups were subdivided into more specific groups. Within the Surgery group, five clinical specialty groups were selected, which included hepatobiliary surgery, thoracic surgery, orthopedics, urology, and neurosurgery. In the internal medicine group, five clinical specialty groups were identified, namely endocrinology, respiratory medicine, gastroenterology, nephrology, and neurology. This resulted in a total of 12 clinical specialty groups. The comparisons of recognition indicators were conducted across these clinical specialty groups.

### Correlation between recognition indicators

The analysis included data from an overall group, as well as segmented data from 8 hospitals and 12 clinical specialty groups. The correlation analysis delved into the interrelationships among main recognition indicators, focusing on four critical aspects: the link between TRP and OAR to assess if there’s a relationship between the extent of recognition and the frequency of overlooked access incidents; the potential linear relationship between TRP and the CHRP; and the correlation between OAR and CHRP. These analyses collectively aimed to provide a comprehensive understanding of how recognition practices, cost implications, and overlooked access interacted within and across hospitals.

### Comparison between specialized and general hospitals

This analysis involved a comparative analysis of recognition indicators on the MR platform between specialized hospitals and their corresponding specialties within general hospitals. Specifically, the study collected and compared data from two distinct groups: Data from the children’s specialized hospital (Hospital H) was gathered and contrasted with data from pediatric departments in general hospitals (excluding Hospital H). The comparison included a range of indicators on the MR platform to assess differences in recognition practices and outcomes between the specialized hospital and general hospital focused on pediatric care.

Similarly, data from the traditional Chinese medicine hospital (Hospital E) was compared with data from TCM departments within general hospitals (excluding Hospital E). This comparison aimed to identify any significant disparities in recognition behavior and to evaluate the effectiveness of recognition within the context of TCM practices across different types of hospitals.

### Comparison between clinical specialties in different hospitals

Hospital management strategies on recognition had an affect on physicians’ behaviors, for example, physician training, performance incentives, and MR promotion, etc. Two large general hospitals with similar scales, Hospitals C and D, both with complete departments of urology, thoracic surgery, orthopedics, nephrology, gastroenterology, and respiratory medicine, were selected as representative cases. The recognition indicators of the aforementioned clinical specialties in the two hospitals were compared to analyze the physicians’ behaviors of the same clinical specialties in different hospital management.

### Information intervention

According to the design of the MR platform, every item that had been accessed must be subject to a recognition decision. However, before reaching the decision-making phase, particularly during the report access stage, the platform did not uniformly require all hospitals to implement information intervention on the dotted arrow path that overlooked access in Fig 1, restricting physician from using this path. When hospitals were connected to the MR platform, their internal information intervention strategies were developed based on the clinical habits of physicians. Ultimately, Hospitals A, C, and F carried out the information intervention on path of overlooked access while other hospitals had not set up. We conducted a follow-up observation for over a year on the recognition behavior of physicians in each hospitals and compared the recognition indicators between hospitals with and without information intervention on path of overlooked access.

This study, which involves the analysis of existing, anonymized data, did not require direct patient or public involvement in data collection. However, we acknowledge the importance of patient and public perspectives and have consulted with relevant stakeholders to ensure our research questions and outcomes are aligned with their interests and needs. Though not directly involved in this observational study, their insights have informed the broader implications and potential applications of our findings. We remain committed to engaging patients and the public in our research endeavors moving forward.

## Results

### Overall and Grouped Results

During the study period, the 8 large hospitals received a total of 26,759,077 delivered items through the MR platform, with 3,639,368 precision delivered items sent to meet physicians’ orders. There were 3,265,038 instances of physicians accessing reports, and 2,449,006 instances where physicians explicitly made recognition decisions. There were 1,190,431 instances where reports were overlooked and not accessed.

The recognition indicators for different clinical specialty groups were displayed in Table 1. There were significant differences in the PDR among the four major clinical specialty groups. The TCM group had the PDR of only 0.76%, which was closely related to the relatively low demand for routine exams and tests and lower attention to past items by TCM physicians. In contrast, the pediatric group had a rate as high as 5.79%, significantly higher than both the surgery group and the internal medicine group. Without distinguishing between specialties and time periods, the TRP within 30 days was 41.54%, with the surgery group and the internal medicine group being roughly the same, while the TCM group had the highest TRP, reaching 51.18%, especially for reports from other hospitals, reflecting a higher recognition of patients’ past reports in TCM diagnostic and treatment concepts. In comparison, the TRP of the pediatric group was significantly lower than other groups, at only 9.95%, indicating that pediatric physicians, due to the rapid changes in children’s conditions, payed more attention to recent item results, and most past items of children had not been recognized.

**Table 1.** Recognition indicators across clinical specialty groups in the MR platform.

| Groups | Clinical Specialty | TRP(%) | CHRP(%) | PDR (%) | REFP(%) | OAR(%) |
| --- | --- | --- | --- | --- | --- | --- |
| Surgery Group | Hepatobiliary surgery | 32.34 | 41.28 | 3.84 | 74.96 | 13.09 |
|  | Orthopedics | 31.05 | 51.63 | 4.81 | 89.04 | 9.52 |
|  | Urology | 21.15 | 24.98 | 9.78 | 28.16 | 16.68 |
|  | Thoracic surgery | 50.67 | 44.44 | 17.77 | 70.23 | 55.65 |
|  | Neurosurgery | 33.28 | 39.14 | 2.85 | 79.76 | 15.46 |
| Internal Medicine Group | Endocrinology | 35.84 | 39.23 | 25.27 | 1.74 | 29.55 |
|  | Neurology | 40.45 | 49.75 | 8.84 | 34.15 | 18.5 |
|  | Nephrology | 69.03 | 57.92 | 42.29 | 1.40 | 57.34 |
|  | Gastroenterology | 44.66 | 52.21 | 6.64 | 16.44 | 14.42 |
|  | Respiratory Medicine | 36.39 | 40.21 | 10.94 | 64.66 | 28.27 |
| TCM Group | TCM | 51.18 | 67.5 | 3.18 | 28.7 | 13.29 |
| Pediatrics Group | Pediatrics | 9.95 | 12.32 | 19.79 | 18.46 | 14.76 |
| All Data |  | 41.54 | 39.81 | 13.6 | 27.31 | 32.71 |

Distinct differences in recognition indicators among clinical specialty groups were shown in Table 1. For instance, the nephrology group had a TRP as high as 69.03%, while the urology group was relatively lower, at only 21.15%. The PDR in the Endocrinology group was 25.27%, while the Neurosurgery group was only 2.85%. These differences reflected the varying degrees of demand and reliance on exams and tests items in different clinical specialty groups during the diagnostic and treatment process, thereby affecting their behavior on the MR platform.

Surgical specialty groups, such as orthopedics, hepatobiliary surgery, and neurosurgery primarily focused on radiological exams, resulting in a relatively higher REFP. In contrast, The internal medicine specialty groups majorly focused on laboratory test recognition, reflecting the characteristic of internal medicine that emphasizes laboratory index analysis. However, the respiratory medicine group payed more attention to radiological exams, leading to a slightly higher REFP compared to other internal medicine specialties. The urology group, due to the high frequency of routine urine tests, had a significantly lower REFP to other surgical specialties.

The TRP, CHRR, and AR were main recognition indicators of the recognition effectiveness across the selected hospitals, which were shown in Table 2. The highest rates in both TRP(75.77%) and CHRP(48.96%) were shown in Hospital D, indicating strong policy adherence and intra-hospital collaboration. Conversely, Hospital H had the lowest rates, suggesting potential barriers to implementation. The AR were generally high in Hospitals A, C, and F. However, The lower rate of 44.61% in Hospital B indicated a need for improvement. The OAR was notably high in Hospital B (55.39%), suggesting possible resistance or lack of awareness, while Hospitals A, C, and F showed low rates, below 2%, related to the information intervention in report access conducted by the three hospitals.

**Table 2:**
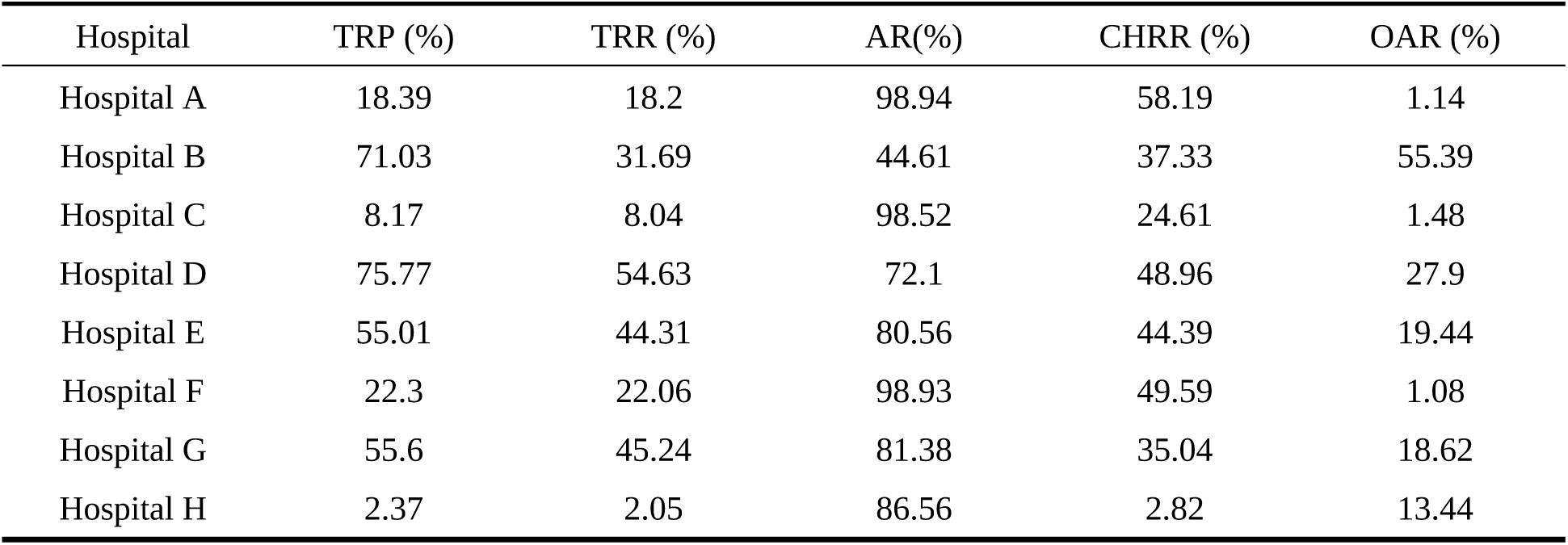
The recognition indicators for selected tertiary hospitals in the MR platform.

### Relationship between major recognition indicators

The regression analysis of three key recognition indicators: OAR, TRP, and CHRP were shown in Fig 2. The 21 data points in each chart are derived from an overall group, as well as segmented data from 8 hospitals and 12 clinical specialty groups. Fig 2A shows the correlation between OAR and TRP, presenting a positive correlation (r=0.682, p<0.05). The correlation indicates that as the rate of overlooked access increased, the rate of total recognition proportion also increased. Fig 2B shows the correlation between CHRP and TRP, presenting a positive correlation (r=0.776, p<0.05). The correlation indicates that hospitals with a higher proportion of cross-hospital recognition also tended to have a higher total recognition proportion. Fig 2C shows the correlation between OAR and CHRP, presenting a non-significant correlation (r=0.268, p>0.05). The positive correlation observed between the OAR and TRP implies that physicians with a higher rate of overlooking to access reports also exhibit a higher TRP. This correlation may stem from the clinical practice where reports overlooked by physicians were typically non-recognizable, thus when the OAR is high, the denominator in the TRP calculation is reduced, which in turn boosts the TRP. The absence of a correlation between OAR and CHRP suggests that the tendency of physicians to overlook delivered reports does not influence their recognition of delivered reports from other hospitals, as the majority of the overlooked reports are likely internal to their own hospital.

**Fig 2.**
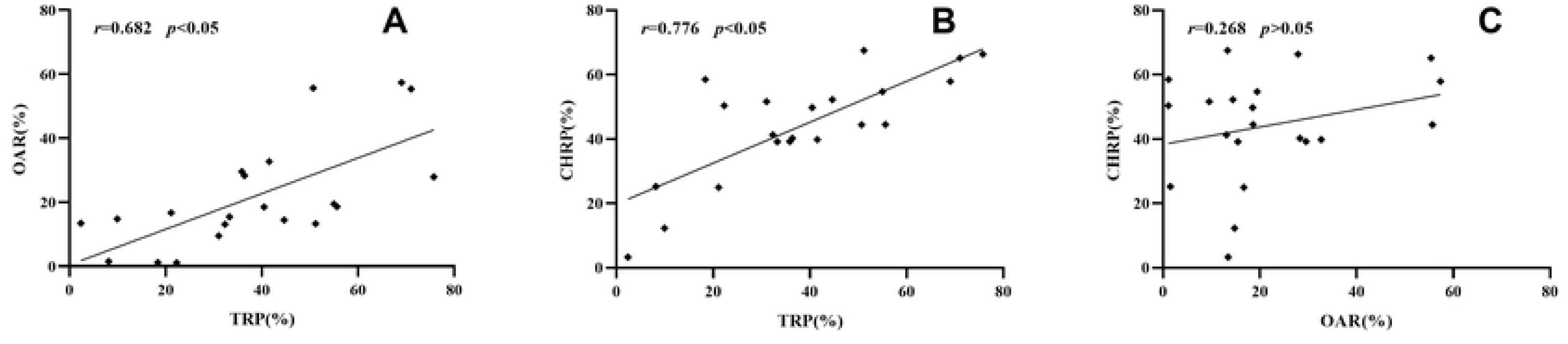
Correlatio analysis of OAR, TRP, and CHR

### Comparison between specialized and general Hospitals

Hospital H represented data from a large pediatric specialty hospital, while the pediatric department data excluded Hospital H and reflected the characteristics of pediatric departments in general hospitals. The recognition indicators reflected physicians’ behavior in terms of disease diagnosis and treatment needs are shown in Fig 3. The figure presents a comparative bar chart of seven recognition indicators for pediatrics in general hospitals (blue bars) and pediatric specialized hospitals (red bars) in Fig 3A, and for TCM in general hospitals (green bars) and TCM specialized hospitals (yellow bars) in Fig 3B. The x-axis represents the rate of each indicator as a percentage, while the y-axis lists the seven indicators. The four indicators, including CHAR, AR, CHRP and PDR, demonstrate no substantial differences between general and specialized hospitals in both pediatrics(Fig 3A) and TCM(Fig 3B). However, the significant disparities are observed in CHRR, TRR and TRP between the two hospitals in Fig 3A. The corresponding indicators for hospital H are significantly lower than those for the pediatric departments in general hospitals, indicating that pediatric specialized hospital physicians have a higher requirement for recent medical reports compared to pediatricians in general hospitals. There are minimal differences in the recognition indicators between general hospitals and TCM specialized hospitals. The CHRR, TRR, and TRP are relatively consistent, indicating that the level of recognition practice is comparable. The differences between TCM departments in general hospitals and hospital E are significantly less pronounced than those between pediatric departments in general hospitals and hospital H. This suggests that TCM has a relatively high recognition radio for recent medical records, and despite being in different hospital settings, the demand differences for recognition do not significantly across TCM specialties.

**Fig 3.**
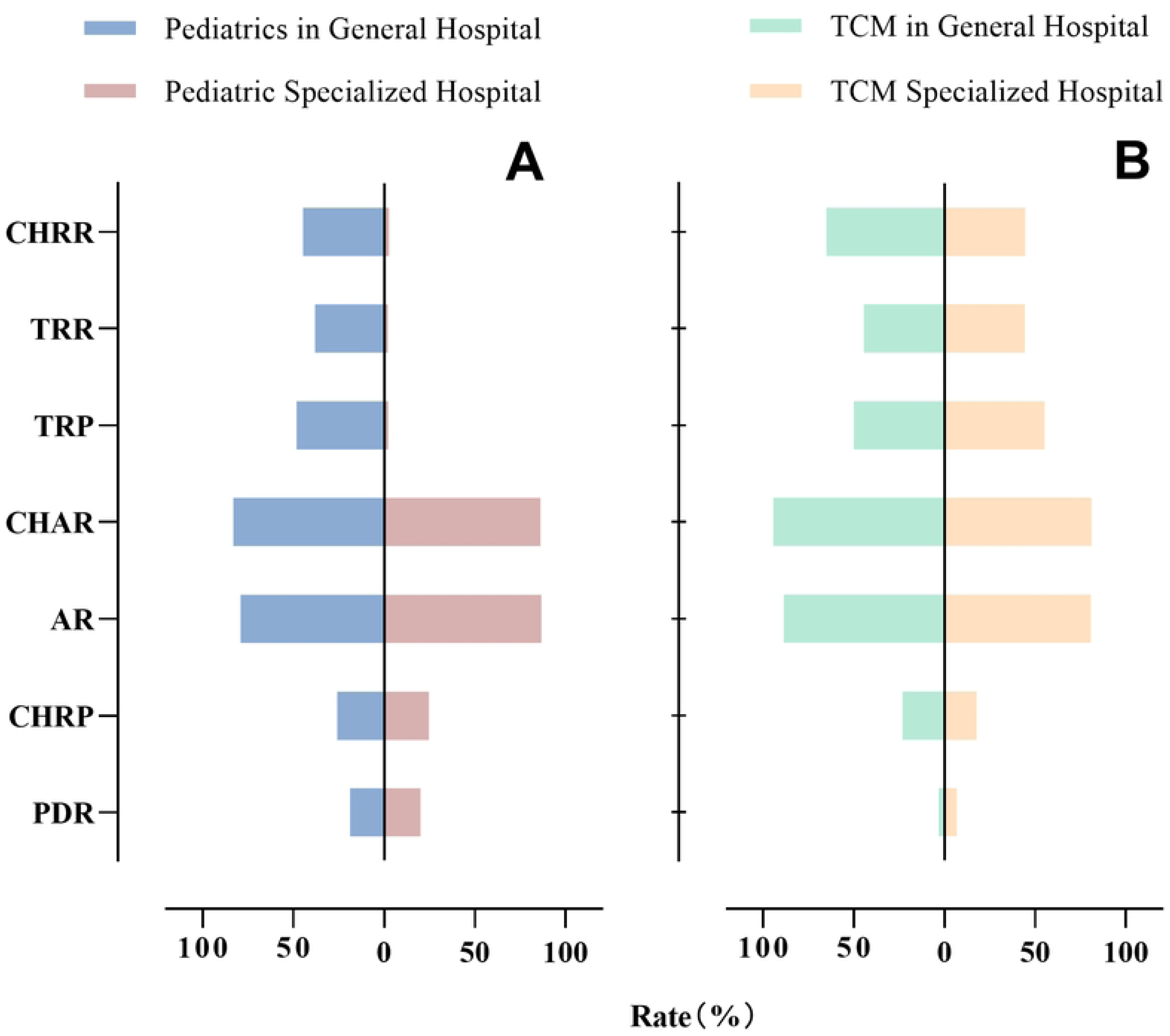
Comparative analysis of seven recognition indicators betw

### Affect of information intervention

It is shown that the implementation of information intervention strategies can significantly reduce the OAR, ensuring that reports delivered by the MR platform are accessed by physicians(Fig 4). Hospitals A, C, and F have implemented information intervention strategies. Under the information intervention strategies, the OAR among physicians have significantly decreased, with rates as low as 1.14%, 1.48%, and 1.08%, respectively. However, the TRP in these three hospitals are notably lower compared to other hospitals without the information intervention strategies, standing at 18.39%, 8.17%, and 22.30%, respectively. Further analysis of the CHRP reveals no significant difference between hospitals with information intervention and those without. This suggests that while the information intervention could effectively reduce instances of overlooked accession to delivered reports, it may also somewhat reduce the TRP of the hospital, although the affect on the CHRP is not significant. It is reflected that there is no significant difference in the recognition of reports to external hospitals due to the intervention, but primarily affects the accession of reports originating from intra-hospital. It is noteworthy that hospital H, a pediatric specialized hospital, despite not having implemented information intervention, has very low TRP and CHRP, which may be related to the special diagnostic and treatment needs and patient population characteristics of pediatric specialized hospitals. It is also reflected that beyond the influence of information intervention, the characteristics of the clinical specialty and other management strategies of the hospital also play an important roles. Consequently, even with an increased OAR, the TRP and CHRP remained relatively low.

**Fig 4.**
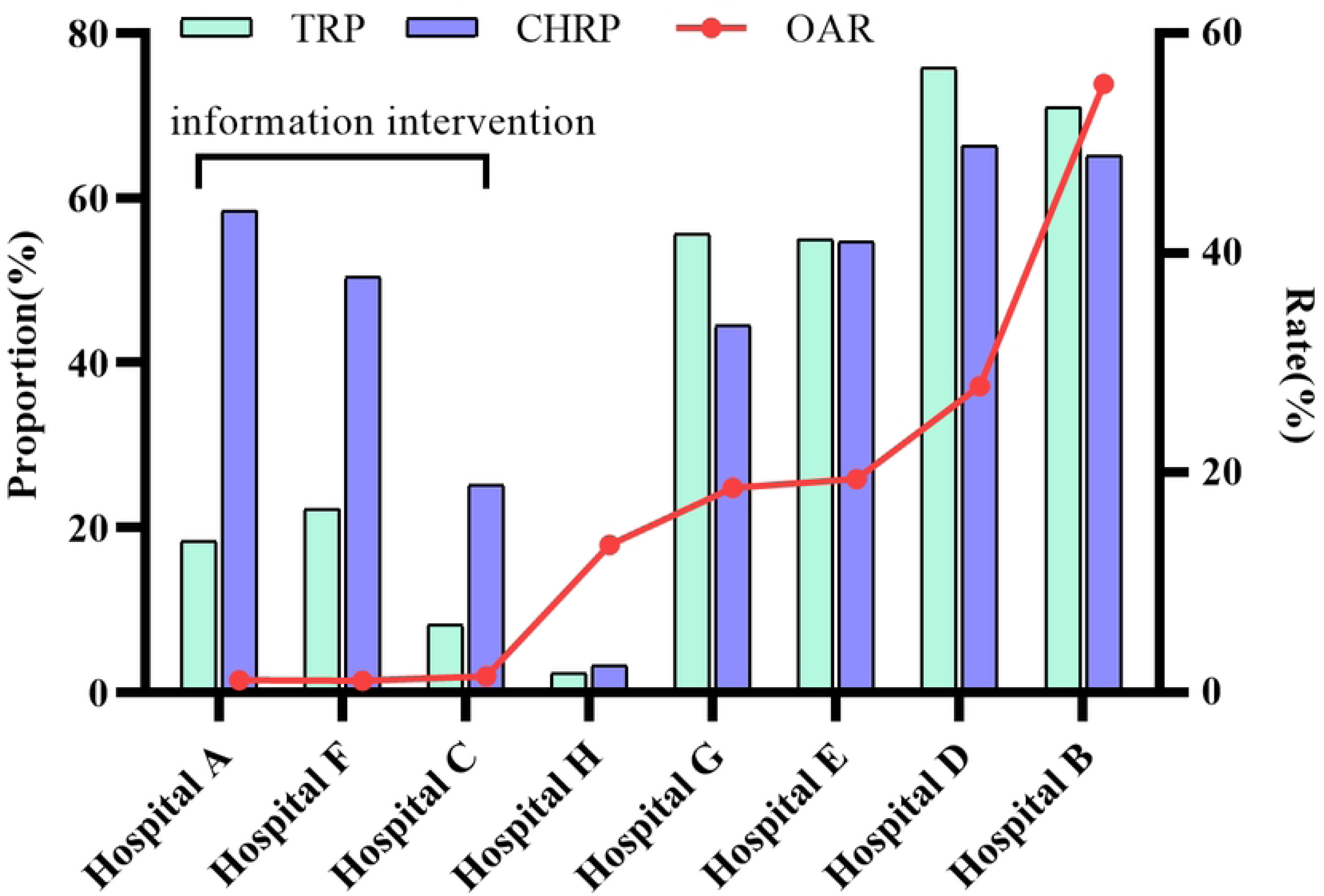
Recognition indicators across various hospitals with inform

### Clinical specialties in different hospitals

A visual representation of the recognition indicators for various clinical specialties in hospital C and hospital D are shown in Fig 5. The indicators include TRR, CHRR, IHRP, CHRP and OAR. The color gradient from light to dark blue signifies increasing values, with darker shades indicating higher rates or proportions.

**Fig 5.**
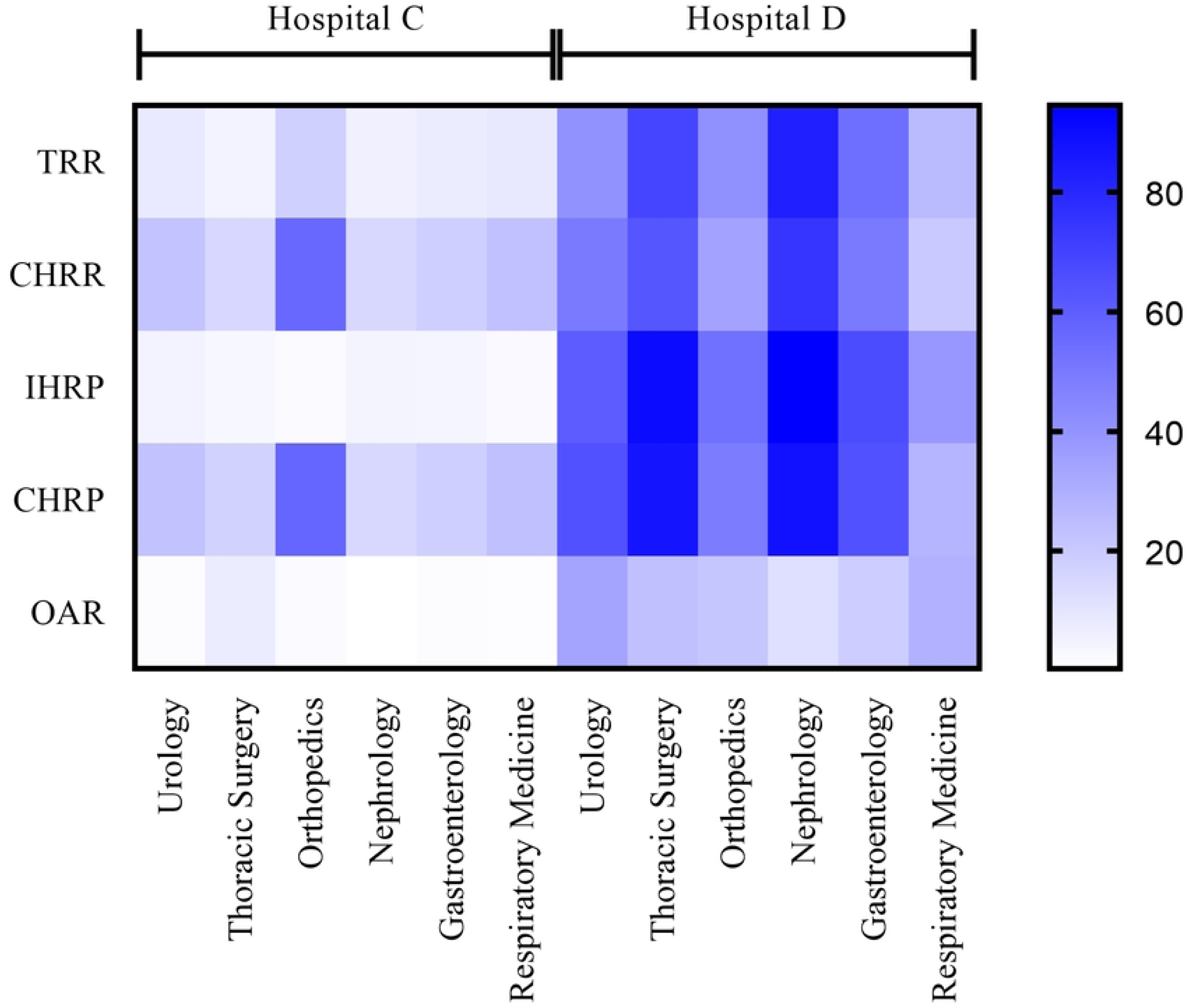
Comparative analysis of recongnition indicators across clini

Hospital D exhibits a generally higher recognition across most indicators compared to hospital C. This is evident from the darker blue shades prevalent in columns of hospital D for TRR, IHRP and OAR, suggesting a more effective implementation of recognition practices. This indicates that while both hospitals may have comparable practices in acknowledging reports from other hospitals.

It is revealed that for hospital C, the TRR, IHRP and OAR for most clinical specialties are generally lower than corresponding clinical specialties in hospital D, indicating that the physicians’ behaviors are significantly influenced by the management of its host hospital. Due to the implementation of information intervention strategies in hospital C, physicians have accessed a higher number of delivered reports that may not require attention, leading to a lower TRR in the final count. However, it is noteworthy that the orthopedics department in hospital C has a higher CHRP compared to hospital D. Orthopedics physicians has a higher recognition for recent radiology reports, and the implement of information intervention in hospital C has promoted an increase in the number of recognition in orthopedics.

## Discussion

### The relationship of MR and DRG

The implementation of DRG reform in China’s healthcare system has significantly influenced the necessity and affect of MR. As highlighted in the study, the DRG reform has led to a notable decrease in the standard deviation (SD) of hospitalization expenditure for patients with chronic obstructive pulmonary disease (COPD), acute myocardial infarction (AMI), and cerebral infarction (CI) in a Chinese city[13,14]. The MR has been catalyzed by the DRG payment reform in China, positively influencing its implementation and leading to a more efficient and patient-centered healthcare model. This integrated approach is crucial for the future of healthcare management and policy-making in China, as it tackles the challenges of escalating healthcare costs and the demand for high-quality, cost-effective medical services.

In the advancement of MR, the recognition needs and scales of clinical specialties are closely related to the main diseases treated, which are main factors influencing recognition practices. By conducting an in-depth analysis of recognition indicators from various large hospitals, this study provides reliable evidence and strong guidance for precisely identifying differences in the performance of MR across different clinical specialties[15]. Since the promulgation of MR by the National Health Commission in 2022[16], a clear standard has been established, strictly following the framework of mutual recognition at the same level. In China’s medical reform, tiered diagnosis and treatment have a significant affect on the use of medical insurance funds and medical services for various diseases.[17,18] Due to the complexity of medical technology, the challenging nature of patient conditions, and the high demand for diagnostic accuracy, theoretically, the recognition rates of large tertiary hospitals may be relatively lower compared to primary and some secondary hospitals. This phenomenon does not imply that the MR work in large tertiary hospitals is lagging; rather, it reflects the need for a more cautious balance between the feasibility and necessity of recognition when facing complex conditions, to ensure medical quality and patient safety. On the other hand, the equipment and quality control levels for exams and tests in lower-level hospitals usually do not meet the standards required by tertiary hospitals. Large tertiary hospitals, with their extensive range of diagnoses and treatments, involve a vast array of diseases, and the indicators related to the recognition practices can comprehensively and meticulously reflect the needs of each clinical specialty for various diseases[19]. Therefore, these hospitals have the capability to set precise reference baselines for recognition indicators within the provincial medical field, providing a clear and definite direction for hospitals at all levels to carry out comparative management work. In this study, the eight large tertiary general hospitals across the province, including general hospitals and various types of specialized hospitals, have been selected, demonstrating good representation and depth in terms of categories, which can accurately and effectively present the behavioral patterns and intrinsic needs of clinical specialties in MR. This ensures that the research conclusions are highly representative and universally applicable, providing crucial and solid support for the optimization of medical resource allocation and the refinement of MR policies.

The recognition indicators constructed in this study offers significant analytical and evaluative value for the recognition behaviors of physicians on the MR platform. Through these indicators, a comprehensive and systematic reflection of the performance of hospitals, clinical specialties, and physicians on the MR platform can be achieved. These indicators provide a quantitative basis for in-depth research on the operational effectiveness of the MR platform, assisting hospital management in accurately assessing the efficiency of MR work, identifying existing issues, and taking timely targeted improvement measures. At the same time, they provide a unified standard for horizontal comparisons between different hospitals, promoting the exchange of experiences and mutual learning among hospitals.

### Optimization of the recognition management in clinical specialties

The needs for medical exams and tests vary significantly among different clinical specialties, which directly affects their behavior patterns on the MR platform. Most clinical specialties have a higher recognition proportion for reports from other hospitals compared to their own hospital reports. This phenomenon is due to various reasons. In the tiered diagnosis and treatment process[20], patients are referred from primary-level facilities to higher-level hospitals. They are usually at the initial stage of consultation, and physicians tend to adopt existing exams and tests reports from other hospitals to quickly establish a diagnosis and formulate treatment plans, saving time and costs for patients, and starting treatment as soon as possible. During the follow-up process within the same hospital, the patient’s condition may be in a state of change, and physicians often need to assess the effectiveness of the treatment at this stage, at which previous reports from the same hospital may not meet the current medical needs, leading to a relatively lower recognition proportion for intra-hospita reports. From the perspective of the recognition process, physicians usually access intra-hospital items within the hospital’s information system during routine medical treatment. It is understandable that physicians do not access the reports delivered by MR platform in the absence of information intervention or other hospital management strategies. This may be the main reason for the high overlooked access rate in hospitals without information intervention. However, there may be other reasons, possibly because some physicians are unwilling or afraid to trust the reports from other hospitals. Therefore, strengthening training on recognition, establishing national or provincial standards for the quality of exams and tests, and implementing them robustly are necessary to alleviate physicians’ concerns about reports from other hospitals.

The recognition indicators of a clinical specialty is not only related to the clinical specialty’s own diagnostic and treatment characteristics but is also closely related to the management and guidance provided by the hospital to the clinical specialty. Taking pediatrics and traditional chinese medicine as examples, there is a significant difference in recognition indicators between pediatric departments in general hospitals and pediatric specialized hospitals, indicating that hospitals can guide clinical specialty physicians to better participate in MR work, improve recognition rates, by developing targeted training programs, establishing effective communication mechanisms, and improving incentive measures. At the same time, hospitals can also formulate personalized recognition guidelines based on the characteristics of different clinical specialties, clarifying which items can be mutually recognized under what circumstances, providing physicians with clear operational standards and reducing inconsistencies in recognition caused by individual judgment differences. In addition, strengthening collaboration and communication between different clinical specialties is also crucial, as good specialty collaboration can promote information sharing and improve recognition efficiency, ensuring that patients benefit from the MR throughout the entire diagnostic and treatment process.

### The affect of hospital management and information intervention

The implementation of appropriate information intervention can furnish physicians with a broader range of patient-related information crucial for decision-making. This encompasses details like the indications, contraindications, and the level of accuracy associated with various exams. Consequently, it empowers physicians to make more judicious choices regarding exam items, thereby enhancing the precision of diagnosis and averting the occurrence of both over-exam and under-exam. Typically, information intervention serves as a principal means by which hospitals conduct the management of medical paths and the enforcement of medical norms. Similarly, information intervention is also regarded as a method for the MR management in some hospitals. On one hand, it is necessary to ensure that the MR platform covers all diagnostic and treatment workstations, enabling physicians to access reports from the MR platform smoothly. On the other hand, effective interventions of the recognition process by physicians is of vital importance. If physicians ignore to access the cross-hospital reports, it will waste the resources for data transmission and increase the exam and test costs that could have been saved on medical insurance. Some hospitals have addressed this issue through information intervention strategies, significantly improving the precision access rate and reducing the overlooked access rate. However, the information intervention strategies also bring some negative affects, such as increasing the workload for physicians in accessing duplicate reports. Under strictly information intervention, physicians must access all delivered reports and make recognition decisions, even if some reports are clearly outdated or not subject to recognition by default, which leads to an increase in non-recognition quantity and a decrease in recognition rate. Therefore, when implementing management strategies, hospitals need to strike a balance between ensuring effective recognition of reports and reducing the workload for physicians in accessing duplicate reports, choosing appropriate management methods based on the actual situation, such as considering the overlooked access rate, total recognition rate, and cross-hospital recognition rate in a comprehensive manner.

### Limitations

Although this study had conducted an in-depth analysis of hospital management and clinical specialty behavior on the MR platform, there were certain limitations. First, the data from the MR platform might only represent a major part of the province-wide mutual recognition, but not the entirety, and did not reflect the full picture in Zhejiang Province, as it did not include data from other reporting platforms, such as the cloud image platform.[21] Second, the information intervention strategies were just one of the methods hospitals use to manage internal mutual recognition; there were other management methods that could also affect the recognition indicators of clinical specialties. These included improving the quality of various exams and tests, conducting promotional training, and performance assessments, etc. In addition, the study did not adequately take into account the demand for exams and tests that originates directly from the patients themselves.

### Future Prospects

In response to the limitations of this study, future research can be carried out in the following areas: First, extend the data collection period and increase the participating hospitals to more accurately grasp the long-term trends and patterns of change, providing more forward-looking basis for policy formulation. Second, strengthen the research on the coordination between medical insurance policies and MR practices, conduct in-depth analysis of the affect of medical insurance payment method reforms, reimbursement ratio adjustments, etc., on the recognition behavior of physicians and patients to provide references for the optimization of medical insurance policies.

## Data Availability

If the data are all contained within the manuscript and/or Supporting Information files, enter the following: All relevant data are within the manuscript and its Supporting Information files.

## Acknowledgments

The authors would like to express their sincere gratitude to all the individuals and organizations that contributed to the successful completion of this study.Special thanks go to the participating hospitals for their cooperation and for the valuable insights they provided into the practical aspects of mutual recognition practices.

